# To snooze, schmooze, peruse, booze, or learn new views: a rejected Christmas Issue research study

**DOI:** 10.1101/2022.12.12.22283344

**Authors:** Joshua D. Wallach, Joseph S. Ross

**Author notes:** **Corresponding author:** Joshua D. Wallach, PhD, MS, Assistant Professor, Department of Epidemiology, Rollins School of Public Health, Emory University, 1518 Clifton Road, Atlanta, Georgia, Room 3028, **Twitter:** @JoshuaDWallach.

## Abstract

**Objective:** To evaluate the frequency of snoozing, schmoozing, perusing, and boozing among attendees at an international conference.

**Design:** Survey and field study

**Setting:** The 9^th^ International Congress on Peer Review and Scientific Publication in Chicago, Illinois (the conference).

**Participants:** 53 attendees (perhaps just enough to be scientific).

**Main outcome measures:** Self-reported conference behavior including frequency of falling asleep (snoozing), interacting with other attendees (schmoozing), and multi-tasking (perusing) during presentations, as well as drinking (boozing) at the conference.

**Results:** 53 attendees completed the survey, which represents a 100% response rate among the attendees who decided to complete the survey. Most of the respondents reported that they preferred in-person conferences before (47/53, 89%) and since (43/53, 81%) the COVID-19 pandemic. 49 (49/53, 92%) reported that multi-day, in-person conferences are still a good use of time. Although 49 (49/53, 92%) reported learning something new (new views), nearly half (24/49, 49%) stated that they could have learned this information from a book or journal article, as opposed to traveling to a conference. Two respondents (2/51, 4%) admitted to snoozing and 43 (84%) to perusing (e.g., emailing, working, and using Twitter) during presentations on the first day of the conference (we suspect that all 5 respondents who claimed that their sleep schedule was “none of our business” also snoozed regularly). The rate of boozing (up to 3 alcoholic beverages) on the first night of the conference was alarming (39/52, 75%), with an additional 10% unable to recall the total number of drinks consumed.

**Conclusions:** Formal, presentation-based conferences may no longer be needed, as attendees appear to be overly preoccupied with snoozing, schmoozing, perusing, and boozing.

**What is already known on this topic:**

- Scientific conferences have functioned as both professional and social events, allowing attendees to learn about the newest research findings while they schmooze, and sometimes booze, with their colleagues.
- Since the COVID-19 pandemic, the research community has grown increasingly accustomed to the benefits of attending conferences from behind their computers.
- Little is known about the behaviors and preferences of attendees following the COVID-19 conference hiatus.

**What this study adds:**

- Almost all respondents reported that multi-day, in-person conferences were still a good use of time and that they had learned something new (even though over half admitted that they could have learned this information from a book or journal article, as opposed to traveling to a conference)
- Conference attendees rarely snoozed but frequently perused (email, work, and Twitter) during presentations; over three-quarters boozed on the first night of the conference.
- Formal, presentation-based conferences may no longer be needed, as attendees appear to be overly preoccupied with snoozing, schmoozing, perusing, and boozing.

## Introduction

Historically, scientific conferences have functioned as both professional and social events,^1^ allowing attendees to learn about the newest research findings (new views, if you will) while they schmooze, and sometimes booze, with their colleagues. Prior to the COVID-19 pandemic, conferences also provided an exciting opportunity for the research community to get out of department meetings, teaching classes, and other responsibilities by traveling away from their bosses, students, significant others, and even perhaps their children. However, soon after the start of the COVID-19 pandemic, in-person conferences were replaced with Zoom-based meetings.

Over the past 2 years, the research community has grown increasingly accustomed to the benefits of attending conferences from behind their computers,^2^ including the capability of nearly undetectable multi-tasking (i.e., perusing) during meetings (including emailing, doom scrolling on Twitter, and online shopping), access to unlimited snacks (without being judged by colleagues), the ability to wear comfortable and casual attire (no pants vs. a tweed suit with elbow patches), and most importantly, the opportunity to alleviate their Zoom fatigue (*ICD-10* code and *DSM-5* definition pending) by snoozing between or during meetings (although folks do this at in-person conferences too). However, as COVID-19 restrictions have eased, in-person conferences have slowly returned.

With limited travel and social interaction over the past few years, a lower tolerance for jet lag, but perhaps a higher tolerance for alcohol, the scientific community is wildly unprepared to re-enter a society with scientific conferencing. In particular, the risk of conference attendees slipping into Zoom-appropriate but real-world inappropriate behavior, such as snoozing or perusing during presentations, has likely increased. Even prior to the pandemic, rigorous and reliable scientific evidence suggested that snoozing during conference presentations was common.^3^ In fact, snoozing was found to be related to presentation duration, dim lighting, warm room temperature, comfortable seating, and time of day.^3^

Currently, little is known about how conference attendees feel about returning to in-person conferences following the COVID-19 hiatus. Although it is possible that conference attendees will be buzzing with excitement about re-engaging in-person with colleagues (i.e., schmoozing and boozing), concerns have also been raised that conferences are no longer useful.^4^ More importantly, we are worried that Zoom fatigue may transform into ‘in-person fatigue’, a concerning new variant, and that conference attendees may now be even more likely to snooze or peruse during presentations. Therefore, we have designed a survey and conducted an informal field study to try to identify the behaviors and preferences of attendees at the 9^th^ International Congress on Peer Review and Scientific Publication in Chicago, Illinois, held from September 8-10, 2022.

Although we submitted our manuscript as an original Christmas Edition research article to the *BMJ*, with what we believed to be a robust sample size, the editors did not rate our work as meeting the research standards of the *BMJ*. We are posting our work on *medRxiv* because we believe that this critical study will help inform whether efforts are needed to fully transition from in-person conferences to metaverse-based conferences, where real-life social interaction and ‘listening’ to presentations will no longer be required. Plus, we hope it will make people laugh!

## Methods

We conducted a survey and field study of attendees at the 9^th^ International Congress on Peer Review and Scientific Publication in Chicago, Illinois (September 8-10, 2022). The aim of the Congress is to “encourage research into the quality and credibility of peer review and scientific publication, to establish the evidence base on which scientists can improve the conduct, reporting, and dissemination of scientific research.” Given that the conference attendees will consist of researchers, journal editors and publishers, members of the media, and others, we believe that this is the perfect population from which to learn about conference behaviors and related views following the COVID-19 hiatus.

### An informal (but highly rigorous) field study

On the first day of the conference, we recorded the (approximate) numbers of attendees and the numbers of attendees that were wearing a mask. To ensure that we were learning new views presented by other researchers, we only measured snoozing and perusing events during the five podium presentations given by members of our research team. Each presentation was 10-minutes long, followed by a 10-minute question and answer session. Snoozing events were defined as any short (e.g., 1 second) or extended forward or backward dropping of any attendees’ head during presentations. Perusing events were defined as any of the following: opening a computer, taking out a smartphone, opening a magazine, newspaper, or book, or any other form of multi-tasking while seated in the auditorium. Perusing events were categorized from afar by trying to look over attendees shoulders. Each author was assigned one half of the auditorium to monitor and logged the number of perusing and snoozing events on the back of used napkins.

### Survey design

The design of our survey was not informed by any previously published surveys, a review of the literature, or discussions with any peers. However, the instrument was pretested by five of our friends, who were warned to keep any legitimate criticisms to themselves. The survey questions were then modified to improve clarity.

To disseminate our survey, we placed a tabletop standing poster (A4 in size) on the display table in the exhibit hall near the entry of the conference on days 2 and 3. We also handed out paper copies of the posters to conference attendees. The poster invited participants to complete the survey and contained instructions encouraging participants to scan a QR code and fill out an online Qualtrics survey. Conference attendees were informed that the survey would take less than 10 minutes. We now acknowledge that this stretched the truth, considering the survey was over 50 questions in length and a probably required closer to 11 minutes to complete. Participation was completely voluntary and anonymous.

### Survey domains

The survey covered 12 main domains, which asked conference attendees about their conference preferences before and after COVID, their views about conferences in general, their experience with ‘Zoom fatigue’, whether they learned anything new at the conference (new views), if they fell asleep during presentations on the first day of the conference (snoozing), how much they interacted with other attendees at the conference (schmoozing), how much they multi-tasked during presentations on the first day of the conference (perusing), how much they drank the first night of the conference (boozing), their travel history to the conference, their accommodation at the conference, their caffeine and food consumption on the second day of the conference, and their personal characteristics.

### Rudimentary Analyses

To ensure reproducibility, we conducted all descriptive statistics by abacus (version 300 Before Common Era). As exploratory analyses, we used bivariate analyses (Fisher’s exact tests) to evaluate which of the survey domains were potential predictors of snoozing or perusing. We then used multi-directional stepwise logistic regression in combination with advanced, state-of-the-art, supervised and unsupervised, big data artificial intelligence machine learning algorithms to select the potentially important variables. Just kidding. Because of limited numbers of survey respondents, we just used the bivariate analyses. Lastly, we created word (well, more like ‘phrase’) clouds, by hand, from the free text responses, because who doesn’t love a word cloud?

It should come as no surprise that the Yale University Institutional Review Board determined this conference survey to pose low risk to individuals volunteering to participate and exempted the survey from full review.

## Patient and Public Involvement

No patients were involved in setting the research question or outcome measures, nor were they involved in developing plans for design or implementation of the study. No patients were asked to advise on interpretation or writing up of results. To be candid, we would have been embarrassed to waste their time when there are more important matters to attend to, like climate change, a war in Ukraine, and the death of the Queen. Instead, for “public” involvement, we shared the survey questions with a handful of our colleagues.

## Results

### Conference Attendees

According to the conference organizers, there were a total of 511 online and in-person conference attendees. Our back of the envelope calculations (# of rows times # of attendees per row) suggests that there were approximately 200 attendees in the main auditorium on each of the 3 days of the conference. From the conference attendee list included in the final program materials, we can confirm that there were attendees from all over the globe, including North America, South America, Europe, Asia, Africa, and Australia/Oceania; to our knowledge, there were no attendees from Antarctica.

### Field Study

Across all 3 days, we observed that approximately 10-20% of the audience was wearing a mask (of the approximately 200 attendees in the main auditorium on each of the 3 days of the conference). During the 5 podium presentations given by our research team, we identified approximately 1-2 snoozing events per presentation. During one of the presentations, one attendee, in the third row from the front, was sound asleep (eyes closed and arms crossed) for the entire presentation.

We observed that approximately half of attendees used their computers or phones during each presentation. Although we cannot claim with certainty that the vast majority of these multi-tasking conference attendees were not just taking notes, we observed a large number of attendees checking their emails, posting or browsing on Twitter, and texting on their phones. We also observed several attendees knitting, looking at Google Images of Queen Elizabeth and Prince Harry, processing manuscript submissions, listening to music, and cleaning their ears with the ends of their glasses.

### Survey

A total of 53 attendees completed the survey, which represents a 100% response rate among the 53 attendees who decided to complete the survey. One-third (18/52, 34%) of the respondents providing demographic data were between 30-40 years of age **(Table 1)**. Investigators (18/52, 35%) and editor/publisher (14/52, 27%) were the most common professions. Although nearly one-third of the respondents (16/51, 31%) were in the entry stage of their careers (<5 years), over one-half (28/51, 55%) were senior (not citizens, per se) professionals with 10+ years experience. There were 16 respondents (16/52, 31%) with podium presentations and 16 (31%) with poster presentations.

**Table 1.** Characteristics of survey respondents, No. (%)

| **Table 1. Characteristics of survey respondents, No. (%)** |  |
| --- | --- |
| **Age group (years)** | **No. = 52** |
| <20 | 0 (0.0) |
| 20-30 | 8 (15.4) |
| 30-40 | 18 (34.6) |
| 40-50 | 8 (15.4) |
| 50-60 | 11 (21.2) |
| 60+ | 7 (13.5) |
| **Profession** | **No. = 52** |
| Editor/publisher | 14 (26.9) |
| Investigator (as in, researcher, not 'private') | 18 (34.6) |
| Editor and investigator | 8 (15.4) |
| Other | 11 (21.2) |
| Retired | 1 (1.9) |
| **Career stage** | **No. = 51** |
| Entry (<5 years) | 16 (31.4) |
| Middle (5-10 years) | 7 (13.7) |
| Senior (10+ years) | 28 (54.9) |
| **Sleep disorder** | **No. = 52** |
| Yes | 2 (3.9) |
| No | 44 (84.6) |
| Should I be telling you this? | 6 (11.5) |
| **Travel time difference** | **No. = 52** |
| Yes | 47 (90.4) |
| No | 5 (9.6) |
| **Caffeine consumption** |  |
| Mean (SD) | 3.2 (3.3) |
| **Breakfast size** | **No = 52** |
| None | 8 (15.4) |
| Small (e.g., fruit) | 12 (23.1) |
| Medium (e.g., yogurt, English muffin, etc.) | 22 (42.3) |
| Large (e.g., English breakfast) | 10 (19.2) |
| 'American' (all you can eat continental/buffet) | 0 (0.0) |
| **Snoozing during day 1 of this conference** | **No. = 51** |
| Yes | 2 (3.9) |
| No, how dare you ask me this! | 44 (86.3) |
| My sleep schedule is none of your business! | 5 (9.8) |
| **Schmoozing during day 1 of this conference** | **No. = 52** |
| None, I sat alone in the corner and ate my lunch in the bathroom stall | 1 (1.9) |
| A few, but less than a dozen (I really only speak to people I have known for years) | 21 (40.4) |
| About a dozen, I will have a conversation with anyone because I have been socially distant for nearly 3 years | 24 (46.2) |
| Dozens, I am pretty popular... | 6 (11.5) |
| **Perusing during day 1 of this conference** | **No. = 51** |
| Yes | 43 (84.3) |
| No | 8 (15.7) |
| **Boozing the night before the conference** | **No. = 52** |
| Yes | 39 (75.0) |
| No | 13 (25.0) |

### Conferencing in general

Most of the respondents (47/53, 89%) reported that they preferred in-person conferences before the COVID-19 pandemic; 43 (81%) reported that they preferred in-person conferences since the start of the pandemic. Three-quarters (40, 75%) of the respondents disclosed that they had attended up to 5 in-person conferences since March 2020; no attendees had attended more than 5 conferences.

Nearly all respondents (49/53, 92%) reported that multi-day, in-person conferences were still a good use of time (and were outraged that we asked this question at this specific conference). Among the respondents who filled out the free-response section, there were various helpful comments describing the future of academic/research conferencing (**Figure 1)**, as well as opinions about characteristics that make conferences exciting (**Figure 2)** and boring (**Figure 3)**.

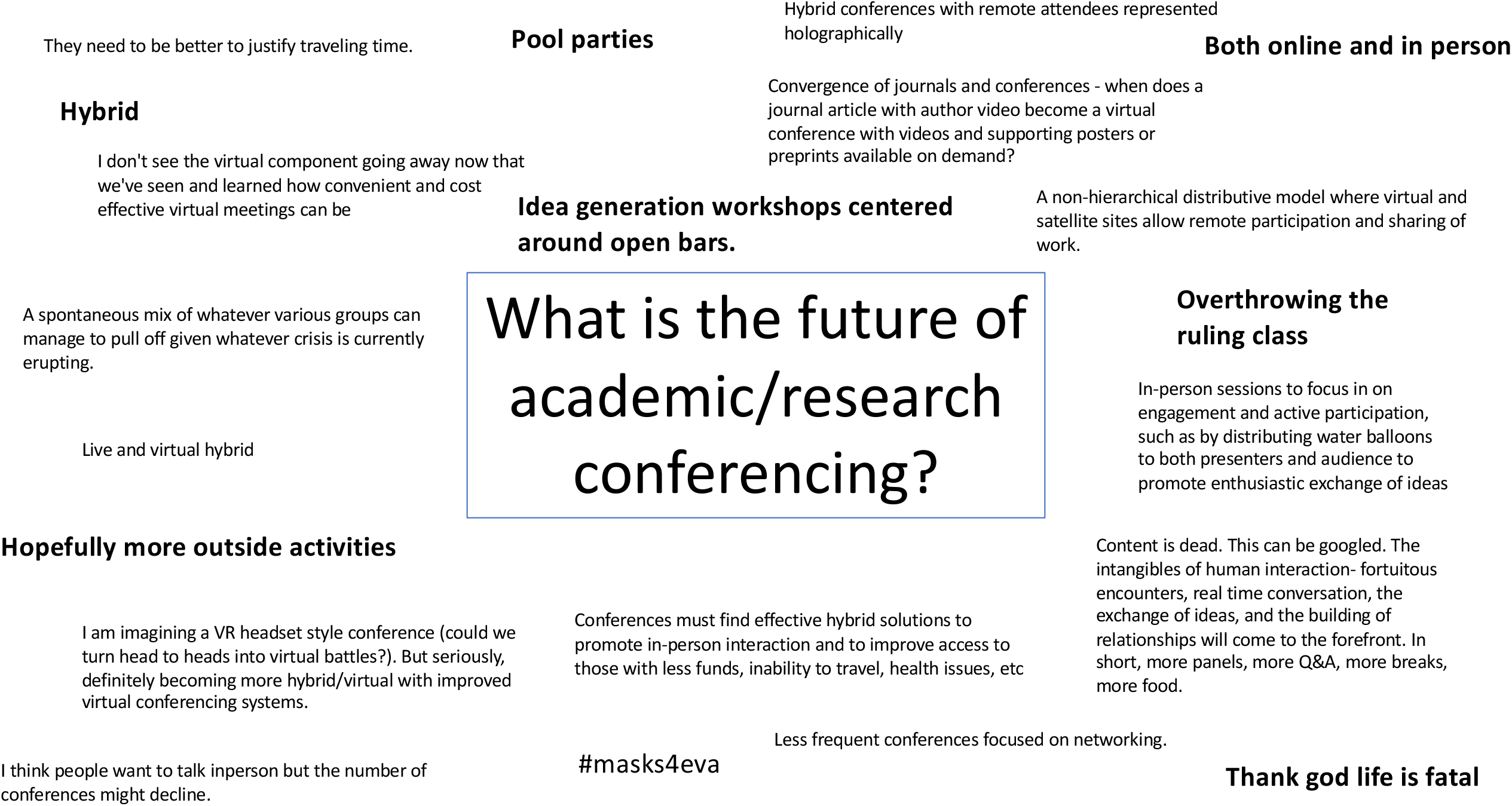

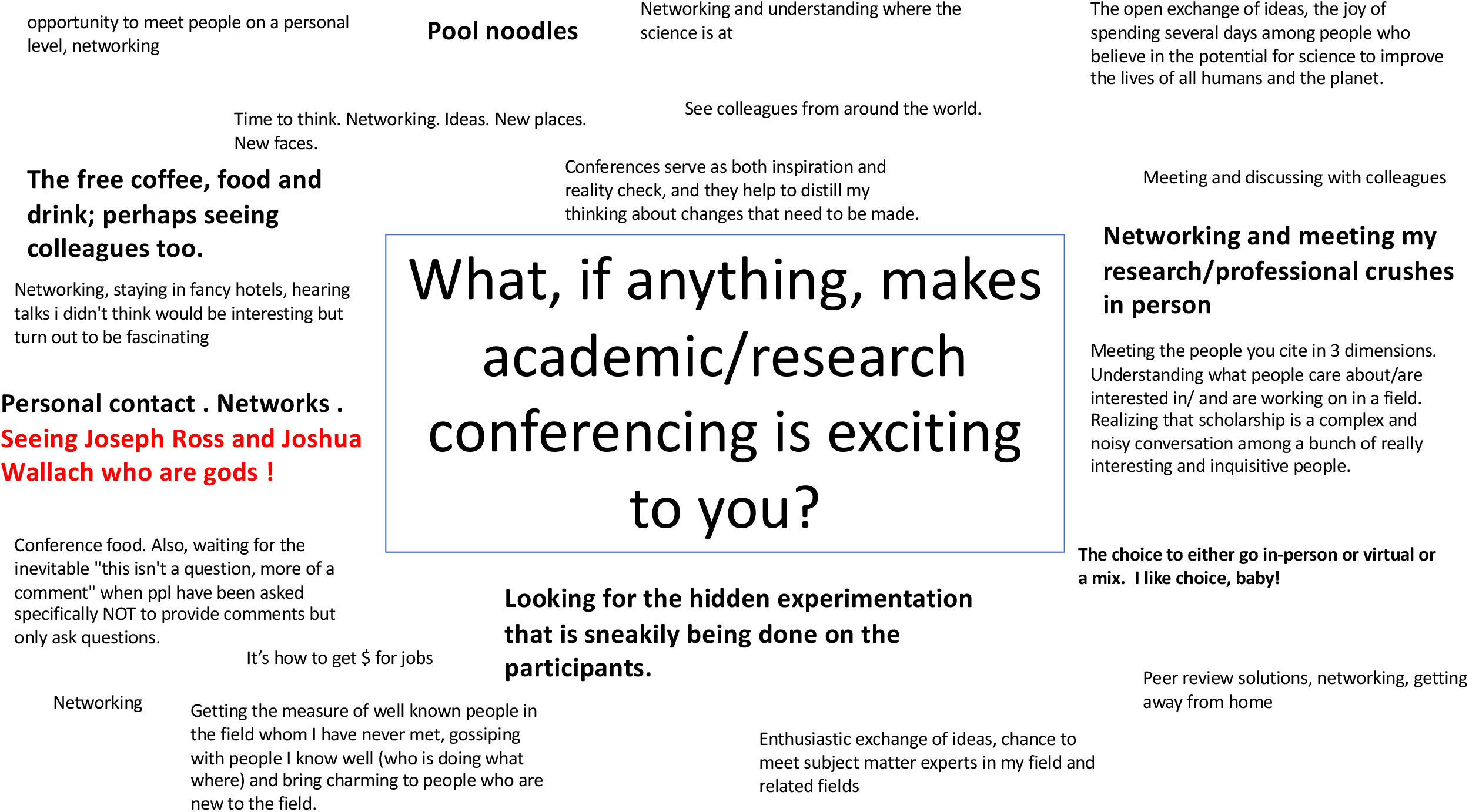

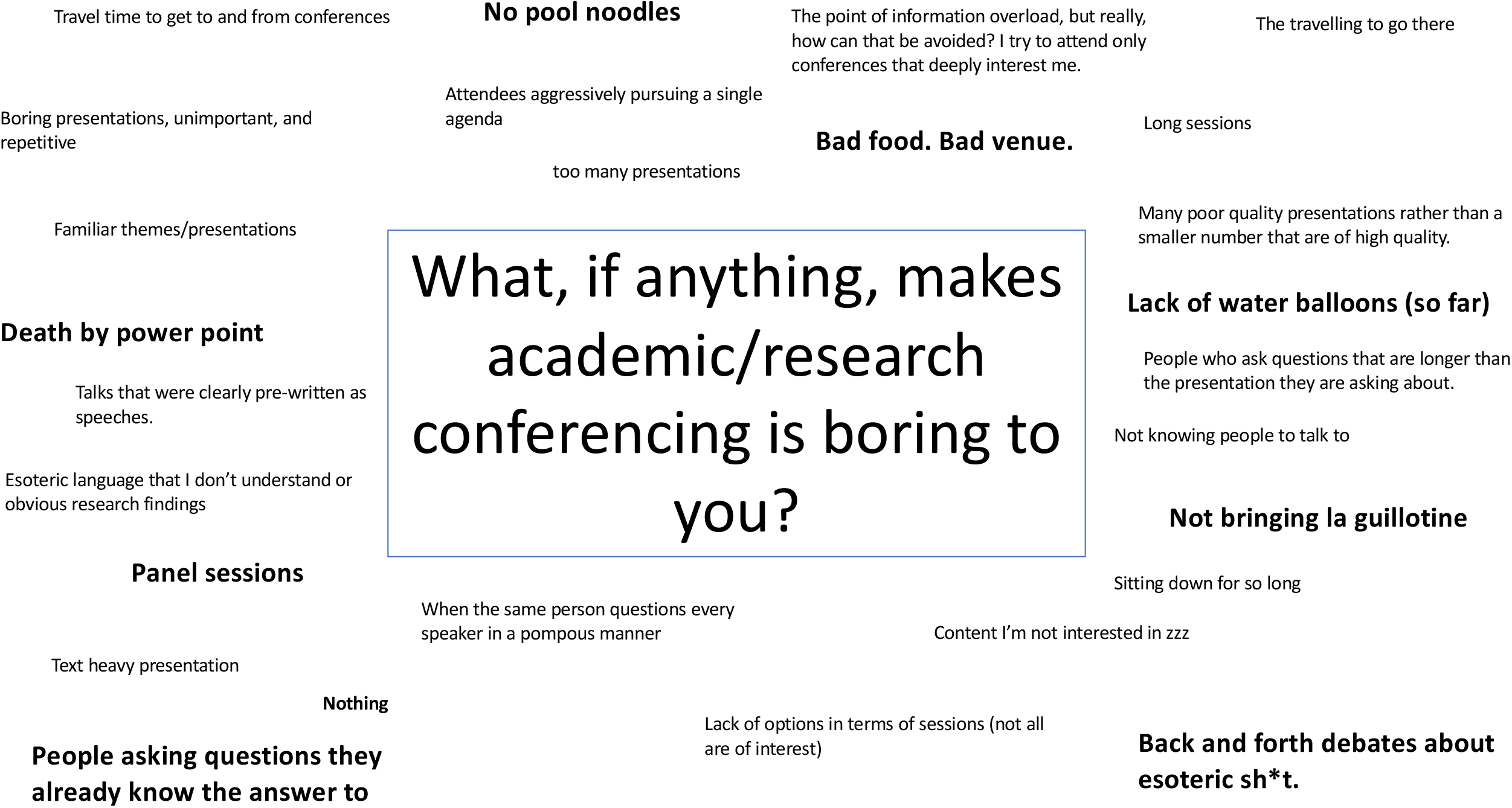

When asked about suffering from Zoom fatigue (on a scale of 0 [not really] to 10 [barely paying attention during Zoom meetings]) and in-person fatigue (0 [not really] to 10 [getting exhausted even thinking about interacting with other humans]), the mean (SD) responses were 7.0 (2.1) and 4.0 (2.3), respectively (N=50). Approximately half (28/53, 53%) reported that their Zoom fatigue was worse than a year prior and one-third (16/53, 30%) reported that their in-person fatigue was worse than a year prior.

### Masking

There were 7 (7/53, 13%) respondents who expected masking to be required at the conference; 37 (70%) expected masking to be optional or not required. Nine (17%) respondents could not believe that we had snuck such a loaded question into our survey.

### New views at the conference

Nearly all (49/53, 92%) of the respondents stated that they had learned something new on the first day of the conference (**Table 1)**. However, when asked to report one thing that they learned, some of the responses were rather concerning: “Queen Elizabeth died”, “Bias sucks”, “The presentations were not as intolerable as I feared”, and “Spinning your articles is good” (**Figure 4)**. Even more concerning, just under half of the respondents who declared that they had learned something (24/49, 49%) stated that they could have learned this information from a book or journal article, as opposed to traveling to a conference.

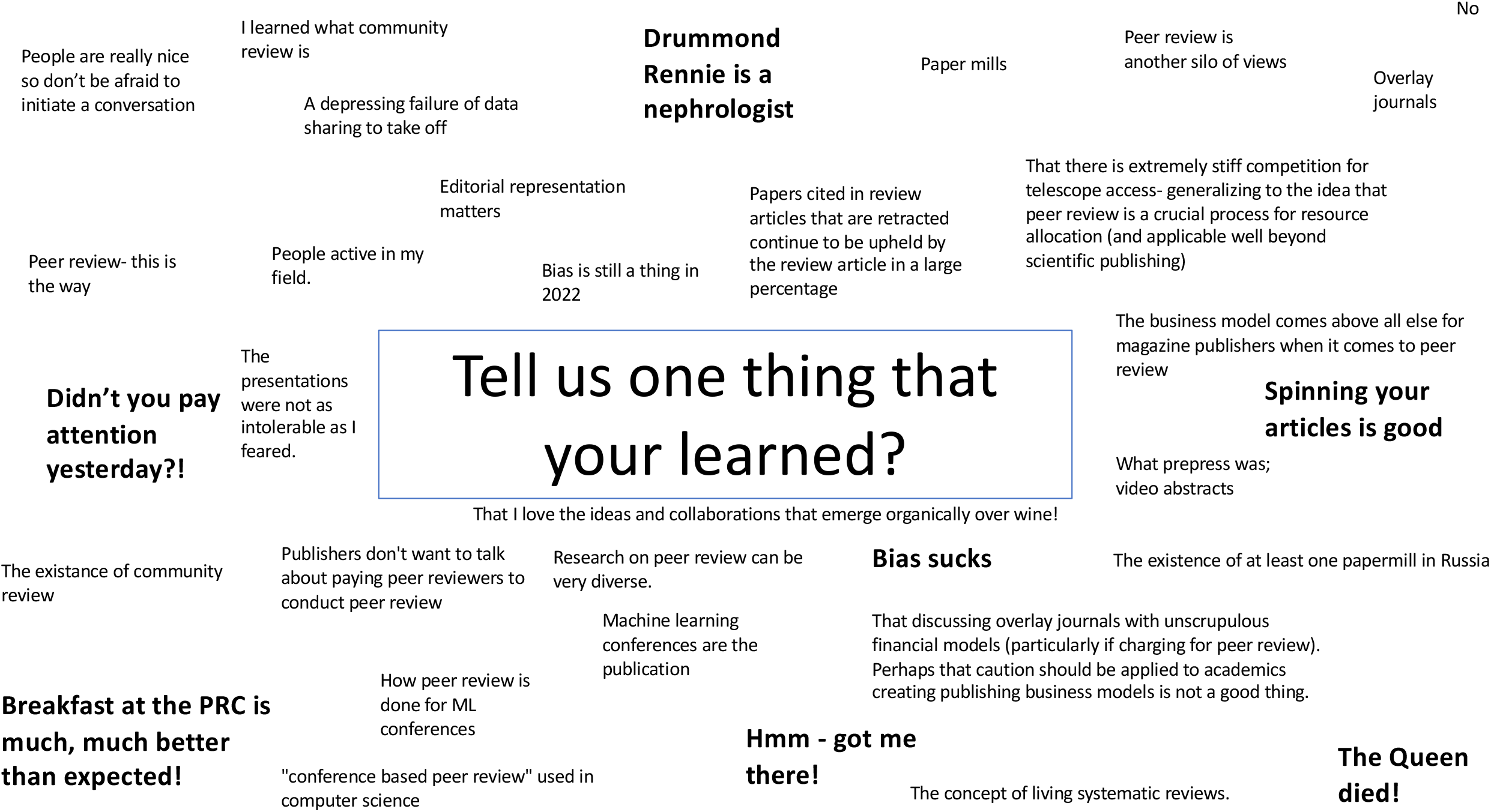

### Snoozing at the conference

Only 2 respondents (2/51, 4%) admitted that they had fallen asleep during day 1 of the conference (**Table 1)**. However, we suspect that most of the 5 respondents (5/51, 10%) who claimed that their sleep schedule was “none of our business” also snoozed regularly. When asked how often they fall asleep at conferences in general, 22 (22/52, 42%) said sometimes and 3 (3/52, 6%) said often (and that conferences are the best cure for insomnia).

Respondents also reported that journal articles (34/52, 65%) and television or books (29/52, 56%) sometimes make them snooze.

### Schmoozing at the conference

Nearly all respondents reported speaking with other people at the conference: 21 (21/52, 40%) claimed that they had spoken to only a few people that they had known for years, 24 (46%) to about a dozen people, and 6 (12%) to dozens of people (**Table 1)**. We were sad to learn that one respondent (2%) sat alone in the corner and ate their lunch in a bathroom stall. Only 19 (19/45, 42%) respondents reported that other people had told them about their experiences feeling isolated during the COVID-19 lock down; 18 (18/45, 40%) had people tell them about their preferences about working from home vs. the office.

### Perusing at the conference

Nearly all (43/51, 84%) of the respondents admitted to multi-tasking on day 1 of the conference (**Table 1)**. When asked about the frequency of reading/writing emails, doing work, or posting/checking Twitter, Facebook or other platforms, 25 (25/52, 48%), 12 (12/52, 23%), and 18 (18/52, 35%) said ‘often’, respectively, but promised that they were great at multi-tasking. All respondents reported having a smart phone (49/49, 100%), of which 41 (84%) said they had used it to Tweet (understandable), whereas 8 (8/52, 15%) actually admitted to shopping online (what a risky decision, considering that the editor in chief of the *BMJ* might have been sitting next to them).

### Boozing at the conference

There were 39 respondents (39/52, 75%) that disclosed that they ‘boozed’ on the first night of the conference (**Table 1)**. While most of these boozers drank in moderation (12 (31%) one drink, 15 (38%) two drinks, and 8 (21%) three drinks – but is that really moderation in a professional setting?), there were 4 (10%) respondents that could not recall the total number of drinks that they had consumed (recall bias or over-indulgence?). Thirty respondents (77%) drank with their colleagues, 5 (13%) drank with strangers, and 4 (9%) drank alone at the bar or in their hotel room.

### Travel, accommodating, and food consumption (i.e., variables potentially related to snoozing during conference presentations)

The mean (SD) travel duration for each respondent was 8.1 (6.7) hours, with most respondents traveling by plane (45/51, 88%) (7 red-eye flights). For nearly all respondents (47/52, 90%), there was a time difference between their home location and Chicago. Unfortunately, 5 respondents (5/51, 10%) had an uncomfortable journey.

There were 41 respondents (41/51, 80%) who stayed at the fancy conference hotel, which received reasonably favorable ratings (out of 10) in terms of bed comfort (mean [SD] 7.9 [1.83]) and water pressure (7.2 [2.2]). Respondents reported a mean (SD) of 6.4 (1.5) hours of sleep the night before taking the survey. Only 2 respondents (2/52, 4%) reported having a known sleep disorder, such as Obstructive Sleep Apnea (with 6 additional respondents questioning if they should actually be answering the question in our survey).

The mean (SD) number of caffeinated beverages consumed on day 1 of the conference was 3.2 (3.3). Most respondents reported having a small (12/52, 23%), median (22/52, 42%), or large (10/52, 19%) breakfast. Fresh fruit, little yogurts, chocolate, and coffee/tea were among the most preferred conference snacks.

### Exploratory bivariate analyses

Lowering the *P*-value threshold for statistical significance to 1.0, we were thrilled to find that nearly all respondent characteristics were associated with snoozing and perusing. There was only one “statistically significant” association between self-reported snoozing and respondent characteristics (a known sleep disorder) and no “statistically significant” associations between self-reported perusing and respondent characteristics using the universally accepted and criticism free *P-*value cut-off value of 0.05. Full results from the bivariate analyses are available upon request (but please, do not bother…).

## Discussion

In our survey and field study of attendees at the 9^th^ International Congress on Peer Review and Scientific Publication, we found low levels of snoozing but high levels of perusing (multi-tasking) during conference presentations. The vast majority of survey respondents reported that they still preferred in-person conferences - likely driven by the desire to schmooze and booze with colleagues - despite disclosing that they could have obtained most of the presented materials from research articles. Lowering the threshold for significance to *P*<1.00, we found that almost all attendee characteristics were “statistically significantly” associated with the likelihood of snoozing and perusing. We believe that the findings from this novel and groundbreaking study should help inform efforts to reimagine, redesign, and improve future conferences following the COVID-19 conference hiatus.

The findings from our survey and field study suggest that approximately 4% of conference attendees snoozed and over 80% perused during podium presentations. Although these numbers may not seem alarming, we remind the reader that many conference attendees consider attending conferences ‘work’. Through this lens, and if one extrapolates these findings to all scientific conferences, the financial implications are immeasurable (‘research waste’^5^). It is also worth noting that nearly half of the respondents were below the age of 30, which means they may be more likely to know how to use personal computers (also referred to as ‘portable desktop computers’) and cellular telephones - the multi-tasking culprits. To mitigate this multi-tasking crisis, it may be necessary for conference organizers to take the following steps: (1) ban all personal computers, Walkman, portable record or LaserDisc players, and cellular telephones and (2) have graded exams following each presentation. To prevent snoozing, conference organizers could also consider implementing higher caffeine consumption requirements (see financial opportunities below).

Most respondents reported that they had boozed and schmoozed with their colleagues. This provides a unique financial opportunity for conference organizers. In addition to the registration and hotel fees, there are lucrative opportunities to charge conference attendees for their alcohol (and snack) consumption. As drinks start to flow, attendees may be more likely to describe their future research ideas, which can then easily be scooped by the conference organizing committees. The members of the organizing committees can then choose to pursue the research ideas themselves or sell the ideas to the highest bidder. Given that conference attendees outlined a desire for more social interaction and outdoor activities (explicitly stating that they did not want to sit as much), conference organizers could further save money by hosting events in public outdoor places and by providing fewer chairs. With survey respondents also admitting that they technically could have learned everything from research articles, formal presentations, with expensive technological support, conferences may no longer be necessary. Moving forward, an outdoor conference with a simple printer (with page charges, of course), a bar (with mark-ups that mirror sporting events, of course), and vending machines selling the most popular snacks, conferences could turn into a profitable enterprise while still ensuring that the needs of conference attendees are fulfilled.

### Challenges conducting survey

When conducting retrospective studies, such as epidemiological surveys, it is important to understand the impact of various biases.^6^ First, researchers must minimize the risk of capturing feedback from only a subset of all potential participants (i.e., sampling bias). We disseminated our survey in the entry hall of the conference, where all participants were required to enter in the morning. If we had only advertised our survey in the main presentation hall, we may have missed the individuals already snoozing after their morning coffee. While it is always possible that participants may have sleepwalked past the announcement in the main hall, it is estimated that <4% of the population suffers from this condition.^7^ To further eliminate any risk of non-response bias, we employed state-of-the-art methodology to increase survey response rates. In particular, we only included dozens of multiple-choice and open-ended questions, and tried different distribution methods (entry hall poster and begging attendees to fill out our survey).

Second, biases can also be introduced if certain participants are systematically more or less likely to recall and relate information depending on their outcome (what academics like to call ‘recall bias’).^8^ Therefore, it is possible that participants who tend to snooze during presentations (and other life events) may have distorted memories about the domains that we surveyed (e.g., flight experience, their breakfast that morning, and the number of drinks they had last night). It is even possible that these participants are remembering their dreams instead of realities. However, as far as we can recall, and assuming we correctly understand how this bias works, recall bias was unlikely to be an issue for our conference survey because only individuals who were awake scanned the QR codes to access the survey. We also surveyed conference attendees about whether they suffered from recall bias while completing the survey and are pleased to report that none of the responders recalled suffering from recall bias (9/52 (17%) definitely not, 13/52 (25%) probably not, and 30/52 (58%) might or might not).

Third, survivorship bias can distort the findings generated by surveys. We are not quite sure about the formal definition of this bias, but we can only assume that it has to do with the number of survey responders who survived the survey questions. We are pleased to inform the reader that unlike many other research surveys, ours was designed to be completely harmless.

Fourth, surveys are also often subject to measurement error. However, we did not take any physical measurements, such as Body Mass Index before and after all the boozing, so we are not sure why this would impact our study. That being said, we felt it was necessary to mention this bias to demonstrate our methodological expertise.

Fifth, we did not use any rewards to motivate survey participation. Although some of the comments submitted by respondents were overly flattering of the authors, it is worth noting that these are real responses. We considered removing these examples from our data but were fearful that this would be considered data manipulation. Sometimes, it is best to just let the data speak for itself. By the time we had reached the conference, we had allocated so much money to registration fees, travel, and drinking alone in our hotel rooms, we literally were not able to bribe respondents to say nice things about us. However, had we used incentives, we would have considered the following prizes to help alleviate snoozing: 1. $100 gift card to Starbucks, 2. A case of Red Bull Energy Drink, 3. A bottle of *Irwin Naturals Brain Awake, Liquid Soft-Gels*, 4. A custom printed travel pillow (“I snoozed at the 9^th^ Congress on Peer Review and Scientific Publication and all I got was this lousy travel pillow”, or 5. One pair of novelty glasses with eyes on them).

Finally, our study’s most serious limitation is that it was rejected by the only journal that could conceivably have published it: the Christmas Issue of the *BMJ*. While the *BMJ*’s decision letter communicated that our survey study did not meet the research standards of the journal, we identified several other Christmas Issue research studies on par with a conference survey of approximately 50 individuals and a 25% response rate, including an ‘n of 1’ study tracking the daily dietary energy intake to maintain a constant body weight of the author during lockdown,^9^ a comparative behavioral analysis of 10 orthopedic trauma surgery experts and 5 Barbary macaques,^10^ and an evaluation of spin in 35 studies on spin.^11^ Thankfully, we had the foresight to launch *medRxiv* in June of 2019,^12^ ensuring that a platform would exist for, what we believe to be, our very clever and funny studies that continue to be rejected by the *BMJ* for the Christmas Issue.^13^

## Conclusions

This survey and field study of attendees at the 9^th^ International Congress on Peer Review and Scientific Publication suggests that there are low levels of snoozing but high levels of perusing (multi-tasking) during conference presentations. Although attendees seemed thrilled about opportunities to schmooze and booze with their colleagues, many admitted that they could have learned information presented from research articles. Formal conferences, with podium presentations, may no longer be needed as attendees only seem to want to booze and schmooze.

## Data Availability

Data are available upon request to the authors.

## Acknowledgements

We would like to thank the organizers of the 9^th^ International Congress on Peer Review and Scientific Publication in Chicago, Illinois for allowing us to disseminate their survey. We are also grateful to the conference attendees who participated in our survey. Lastly, we would like to thank Osman Moneer, Guneet Janda, Xiaoting Shi, and Meera Dhodapkar for their survey feedback and moral support during the conference.

## Contributors

JDW and JSR were responsible for all aspects of the study. All authors participated in the interpretation of the data and critically revised the manuscript for important intellectual content. JDW had full access to all the data in the study and takes responsibility for the integrity of the data and the accuracy of the data analysis. JDW is the guarantor.

## Patient consent

Not required

## Ethical approval

The Institutional Review Board of Yale University determined this study to pose low risk to individuals volunteering to participate and exempted the study from full review.

## Data sharing

The dataset will be made available via a publicly accessible repository on publication.

## Transparency

JDW (manuscripts guarantor) affirms that the manuscript is an honest, accurate, and transparent account of the study being reported; that no important aspects of the study have been omitted; and that any discrepancies from the study as planned (and, if relevant registered) have been explained.

## License

The Corresponding Author has the right to grant on behalf of all authors and does grant on behalf of all authors, a worldwide license to the Publishers and its licensees in perpetuity, in all forms, formats and median (whether known now or created in the future), to i) publish, reproduce, distribute, display and store the Contribution, ii) translate the Contribution into other languages, create adaptations, reprints, include within collections and create summaries, extracts and/or, abstracts of the Contribution, iii) create any other derivative work(s) based on the Contribution, iv) to exploit all subsidiary rights in the Contribution, v) the inclusion of electronic links from the Contribution to third party material where-ever it may be located; and, vi) license any third party to do any or all of the above. The default license, a CC BY NC license, is needed. This is an Open Access article distributed in accordance with the Creative Commons Attribution Non Commercial (CC BY-NC 4.0) license, which permits others to distribute, remix, adapt, build upon this work non-commercially, and license their derivative works on different terms, provided the original work is properly cited and the use is non-commercial. See: http://creativecommons.org/licenses/by-nc/4.0/.

